# Deep metric learning for few-shot X-ray image classification

**DOI:** 10.1101/2023.08.27.23294690

**Authors:** Jakub Prokop, Javier Montalt Tordera, Joanna Jaworek-Korjakowska, Sadegh Mohammadi

**Author notes:** Corresponding author, (JP). These authors contributed equally to this work (Jakub Prokop, Javier Montalt Tordera).

## Abstract

Deep learning models have proven the potential to aid professionals with medical image analysis, including many image classification tasks. However, the scarcity of data in medical imaging poses a significant challenge, as the limited availability of diverse and comprehensive datasets hinders the development and evaluation of accurate and robust imaging algorithms and models. Few-shot learning approaches have emerged as a potential solution to address this issue. In this research, we propose to deploy the Generalized Metric Learning Model for Few-Shot X-ray Image Classification. The model comprises a feature extractor to embed images into a lower-dimensional space and a distance-based classifier for label assignment based on the relative distance of these embeddings. We extensively evaluate the model using various pre-trained convolutional neural networks (CNNs) and vision transformers (ViTs) as feature extractors. We also assess the performance of the commonly used distance-based classifiers in several few-shot settings. Finally, we analyze the potential to adapt the feature encoders to the medical domain with both supervised and self-supervised frameworks. Our model achieves 0.689 AUROC in 2-way 5-shot COVID-19 recognition task when combined with REMEDIS (Robust and Efficient Medical Imaging with Self-supervision) domain-adapted model as feature extractor, and 0.802 AUROC in 2-way 5-shot tuberculosis recognition task with domain-adapted DenseNet-121 model. Moreover, the simplicity and flexibility of our approach allows for easy improvement in the feature, either by incorporating other few-shot methods or new, powerful architectures into the pipeline.

## 1 Introduction

Deep Learning (DL) has shown immense potential in revolutionizing medical image analysis. With access to sufficient data, DL models can achieve human-level performance in a wide range of tasks – from accurate diagnostics comparable to physicians to medical scene perception [1]. However, the main drawback of traditional DL models lies in their heavy reliance on extensive labeled data for effective training on specific tasks. Acquiring and annotating such large datasets can be expensive, especially in the medical domain.

In recent years, few-shot learning (FSL) has emerged as a promising solution to address the limitations of traditional DL models.FSL represents a diverse set of technologies aimed at enabling models to learn and generalize from a limited amount of labeled data, even with novel and unseen tasks. Implementing these technologies holds promising prospects for substantially decreasing the investment necessary for developing novel DL applications through the mitigation of data collection and annotation requirements and the reduction of computational resource demands. Furthermore, FSL techniques can facilitate DL in domains where substantial data availability is lacking, broadening the applicability of DL to various fields.

Radiology is one area with vast potential to benefit from FSL techniques. A frequent use of DL applications in radiology is image classification, closely related to the common radiological interpretative task of providing a diagnosis. Examples include tuberculosis recognition [2], mammographic tumor classification [3], and bone age assessment [4], among many others [5]. These applications can assist radiologists by providing a ”second opinion,” speeding up triage, reducing miss rates, or allowing them to divert attention to more complex cases or tasks. When confronted with the classification of rare or emerging diseases, such as COVID-19 diagnosis [6, 7, 8] and diverse lung pathologies recognition [9, 10], the utilization of the FSL techniques proves invaluable.

Among the FSL approaches, metric learning has emerged as a simple yet highly effective method. Although the idea is long-established, recent results suggest that the metric-based approach remains one of the most powerful FSL methods, outperforming other more sophisticated state-of-the-art algorithms [11]. Metric learning (embedding learning) measures samples’ similarity with defined metrics. It embeds the data samples into a lower-dimensional latent space to reduce the spacing between similar samples and increase the distance between dissimilar ones. Various metric learning approaches have been proposed in the scientific literature to address the problem at hand. Examples of such approaches include *Matching Nets* [12] and its modifications [13, 14, 15, 16], which aim to learn embedding functions for data samples. Additionally, *Prototypical Networks* [17, 18, 19, 20] have been introduced, leveraging the concept of developing class prototypes to enhance metric learning capabilities. Metric learning methods were successfully applied in radiology in tasks such as brain tumor classification [21] and recognition of various chest X-ray pathologies [22].

The recent advance of self-supervised learning [23] further boost the potential of metric learning in medical imaging. Self-supervision can be viewed form of unsupervised learning solving typically supervised tasks. A self-supervised model is either learning to recover missing data parts, e.g. by predicting the masked fragments of an image (generative approach), or learning to predict the similarity of two fragments of the same data sample, e.g. two augmented versions of one image (contrastive approach) [24]. As the self-supervision does not require labelled data, it correlates particularly well with the few-shot paradigm, and perfectly fits the realities of low availability of labelled medical data. Meanwhile, self-supervised models has already proved to be highly effective in many medical tasks [25, 26]. With the flexibility of metric learning approach in terms of model architecture, highly efficient self-supervised models can be easily incorporated into few-shot metric-based pipeline.

A systematic evaluation of different few-shot metric learning models, both supervised and self-supervised, still needs improvement. Metric-based approaches, such as *Prototypical Networks*, allow many variations of model architecture, training strategy and classifier. Yet, to the best of our knowledge, there is no such comparison available for medical imaging. Therefore, we evaluated the method described in [17] on various X-ray classification tasks, implementing several base model variants using more modern architectures and pre-training frameworks.

The main contributions of this paper are as follows:

- We analysed the effectiveness of the few-shot metric learning approach inspired by ProtoNet [17] in radiology, namely in COVID-19 and tuberculosis X-ray classification tasks.
- We benchmarked three convolutional neural network (CNN) and three vision transformer (ViT) architectures as medical feature extractors, as well as three commonly used distance-based classifiers in several few-shot settings, under different data imbalance conditions.
- We compared 6 architectures trained on natural images with 5 domain-adapted ones, including CNN models from TorchXrayVision [27] and RadImageNet [28], as well as with the self-supervised in-domain REMEDIS [29] model based on ResNet-50.
- We fine-tuned DINO-ViT, DINO-ResNet-50 [30] and ViT-MAE models [31] within their respective self-supervised frameworks, using CheXpert [32] X-ray image dataset. We evaluated the effectiveness of this domain adaptation on 11 in-distribution (ID) and 9 out-of-distribution (OOD) disease classification tasks.
- In COVID-19 and tuberculosis classification tasks our model achieved 0.689 and 0.802 AUROC in 2-way 5-shot setting, as well as 0.782 and 0.903 AUROC in 2-way 50-shot scenario.

## 2 Methods

A generalized metric learning model consists of a feature extractor, to embed an image into a lower-dimensional space, and a distance-based classifier, to assign labels to the test samples basing on the relative distance of these embeddings. We followed this schema, and implemented our model as shown in Fig 1. We incorporated several neural network architectures as task-agnostic feature extractors, and described them in detail in Section 2.3. Our selection of classifiers is described in Section 2.4. To evaluate our model in varied few-shot settings, we generated a number of episodes, in which a small, task-specific support set was sampled to fit the classifier (see Section 2.2 for details). Then, the evaluation was performed on the test sets referring to different target tasks, which are given a comprehensive overview in Section 2.6 and Section 2.5.

**Figure 1:**
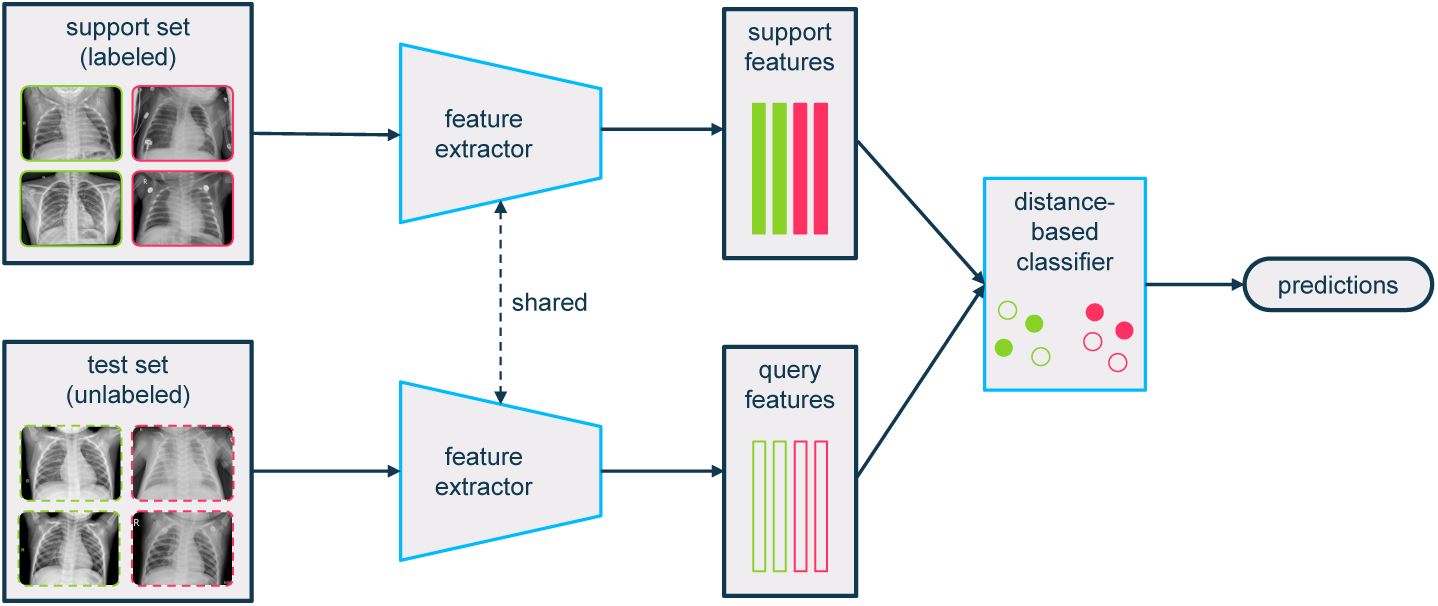
An overview of the metric learning model used in this work. In few-shot learning, the term *support set* denotes a small set of labeled examples used as reference, while the *test set* contains the examples of interest. We used a shared, pretrained feature extractor to capture important information from both sets and obtain encoded feature vectors. These vectors were then passed to a classifier, which predicts the labels of the unlabelled query set. Note that only the classifier is trained with the support set and the feature extractor remains frozen.

### 2.1 Metric learning

The goal of metric learning is to embed each sample ***x_i_*** ∈ *X* ∈ R*^n^* to a lower-dimensional ***z_i_*** ∈ *Z* ∈ R*^m^*, *m* < *n*, such that similar samples are close together and can be easily grouped by a distance-based classifier. In the simplest case, given image-label pairs (***x*_1_**, *y*_1_), (***x*_2_**, *y*_2_), unlabelled image 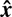 and distance function *d*, the metric learning model would compute ***z*_1_**, ***z*_2_**, 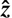, and assign the label *y*_1_ to 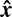 if 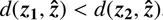, and *y*_2_ to 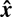 otherwise.

For the purpose of encoding *x_i_* into *z_i_* we utilized a neural network feature extractor, which is trained using task-agnostic supervised or self-supervised learning. To find a correct label *y_i_*, instead of directly measuring the distance *d* to closest labelled sample as described above, we used a distance-based classifier *c* such as k-NN (see Section 2.4). To fit the classifier, we followed [12] and defined a small, task-specific *support set* 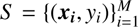, where *M* refers to its size. Then, given a task-specific classifier *c_S_* and a sample from a test set 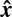, we embedded every ***x_i_***and 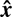 into ***z_i_***, 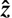 and predicted 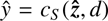 . In our work the distance function *d* is defined as the Euclidean distance, as done in [17].

### 2.2 Few-shot scenarios

The few-shot classification problem is usually referred to as *k*-shot *N*-way classification task [12], where *k* denotes the number of labeled samples for each category in the training set, and *N* refers to the overall number of classes. We implemented four different few-shot settings with *k*∈ {5, 10, 25, 50}.

Our model was assessed using various unseen target tasks, which are described in Section 2.6. Several variations of the model were built with a selection of feature extractors and classification heads. Therefore, we made an evaluation with every combination of *k*-shot setting, target task, feature extractor, and classifier. For each of these combinations, 200 few-shot scenarios (episodes) (*s*_1_, *s*_2_, …, *s*_200_) were randomly generated to reduce random effects and assess the statistical significance of obtained results. For every scenario *s_i_*, a vector ***k_i_*** = (*k_i_*_,1_, *k_i_*_,2_, …, *k_i_*_,*N*_) ∈ N*^N^* was randomly selected, with the value *k_i_*, *_j_* denoting number of examples for class *N_j_* in the support set *S _i_*, under the following constraints:

1. The number of examples for any class in the support set was at least 20% of *k*:

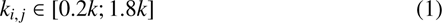
2. The total size of the support set was equal to *k* times *N*:

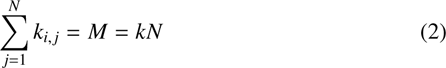

For example, for a 50-shot *2*-way task, there were at least 10 examples for every class and 100 examples in total. This sampling strategy allowed us to study classifier performance under different imbalance ratios. In our experiments we considered only binary classification tasks, giving *N* = 2 and ***k_i_***= (*k_i_*_,1_, *k_i_*_,2_).

### 2.3 Feature extractors

#### 2.3.1 O**ff**-the-shelf models

Our study evaluated the performance of widely used and publicly available neural networks employed as feature extractors. Since these networks were initially pre-trained on natural images, we designated them as ”general-purpose” feature extractors. The analysis included the following models:

- ResNet-50. ResNet [33] is a deep CNN that uses residual connections to alleviate the vanishing gradient problem. Its 50-layer variant is the most used by far and was also used in this work to facilitate comparison with related literature.
- DenseNet-121. DenseNet [34] is a CNN that incorporates feed-forward connections from each layer to every other layer, improving feature propagation and parameter efficiency. Its 121-layer variant was used in this work as it has become particularly popular within the research community.
- ConvNeXT-XL. ConvNeXT [35] is a more modern convolutional architecture, in which the authors systematically studied many design decisions, inspired by recent advances in vision transformers. The extra-large (XL) variant was used in this work because it offers the best performance on ImageNet.
- DINO-ViT-B/8. DINO-ViT [30] is a ViT trained with a self-supervised method called DINO (self-distillation with no labels). Apart of excellent classification performance, the model performs particularly well when combined with a basic nearest neighbors classifier (k-NN), which resonates with our own selection of classifier heads. The B/8 variant (patch size 8) was used due to its classification accuracy on ImageNet.
- DINOv2-ViT-B/14. DINOv2 [36] is the second release of the DINO framework, which produced models with even higher ability to extract high-performance visual features. The B/14 distilled variant was used.
- ViT-MAE-B. ViT-MAE [31] is a ViT trained using masked autoencoding (MAE), a simple self-supervision method that involves masking and reconstructing a large proportion of the image. The base variant with patch size 16 was used in this work.

For CNNs, the feature vector passed as input to the classification head was the output of the global average pooling layer. For ViTs, the class token was passed.

Besides the general feature extractors, we added five ”domain-specific” models to the pool. ResNet-50 and DenseNet-121 have publicly available weights derived from medical datasets published by RadImageNet (RIN) [28] and TorchXRayVision (XRV) [27]. RIN weights are derived from several medical imaging modalities but do not include X-rays. XRV weights are derived primarily from X-ray images. In total, four models were included: ResNet-50-RIN, ResNet-50-XRV, DenseNet-121-RIN and DenseNet-121-XRV.

Lastly, we incorporate REMEDIS-CXR-50-M model. The ”CXR-50” variant of REMEDIS (”Robust and Efficient Medical Imaging with Self-supervision”) [29] is a ResNet-50 architecture initialized with BiT-M [37] weights from natural domain. It was trained within SimCLR [38] contrastive self-supervised framework using CheXpert [32] and MIMIC-IV-CXR [39] large-scale chest X-ray datasets.

#### 2.3.2 Self-supervised domain adaptation

Several sources ([40, 41, 42, 29, 43]) suggest that self-supervised medical domain-adaptation may result in better performance than the supervised approach. Zhou et al.[42] verifies this hypothesis for the ViT-MAE model, while Matsoukas et al. [41] do the same for DINO. In both cases, the obtained improvements seem promising, though in some cases, marginal. Finally, Azizi et al. [29] report outstanding performance of self-supervised domain-adapted models across many different medical image classification tasks, which further boosts our motivation.

We fine-tuned DINO-ViT-B/8 and ViT-MAE-B within their respective self-supervised frameworks to evaluate this approach. We did the same with ResNet-50 using DINO, as this architecture is often used as a backbone for self-supervised models [44, 30, 38, 29]. The pre-trained models and training code are publicly available for both frameworks^1,2^. For training, we utilized the CheXpert dataset [32], containing 224,316 chest radiographs of 65,240 patients, from which 223,648 images were used as our training set (we followed the default split). In both cases (DINO and MAE), we trained the model for 50 epochs, with five warm-up epochs, on 8 A100 40GB GPU units. The batch size was set to 8 per processing unit, and the rest of the hyperparameters were left at their default values.

### 2.4 Classifiers

The following classification algorithms were included in the analysis:

- k-nearest neighbors (k-NN) assigns each observation to the class most common among its k nearest neighbors. k-NN is the most widespread non-parametric classifier. The number of neighbors participating in the vote was set to the expected number of examples per class [45]. Votes were weighted by the inverse of the distance to the observation, as done in [30].
- Nearest centroid (NC) assigns each observation to the class of the training samples whose mean (centroid) is closest to the observation. NC is a simple classifier that has been successful in few-shot settings [17].
- Neighborhood Components Analysis (NCA) learns an optimal distance metric for the nearest neighbors (k-NN) classifier.

All distances were measured using the Euclidean distance, as done in [17] and [45]. The performance of each classifier was evaluated according to the area under the receiver-operating characteristic curve (AUROC) and balanced accuracy – the arithmetic mean of sensitivity and specificity.

### 2.5 Datasets

We used four chest X-ray datasets in our work. We show samples with exemplary metadata from these datasets on Fig 2. The COVID-19 image data collection project [46] collects images from several sources. The dataset has independent labels for several conditions and is suitable for multi-label classification. To prepare the dataset for binary classification, the label vectors containing the COVID-19 label were set to positive, and those without it were set to negative, resulting in 584 positive and 1841 negative cases. The dataset was split in proportions 80%/20% for train and test subsets.

**Figure 2:**
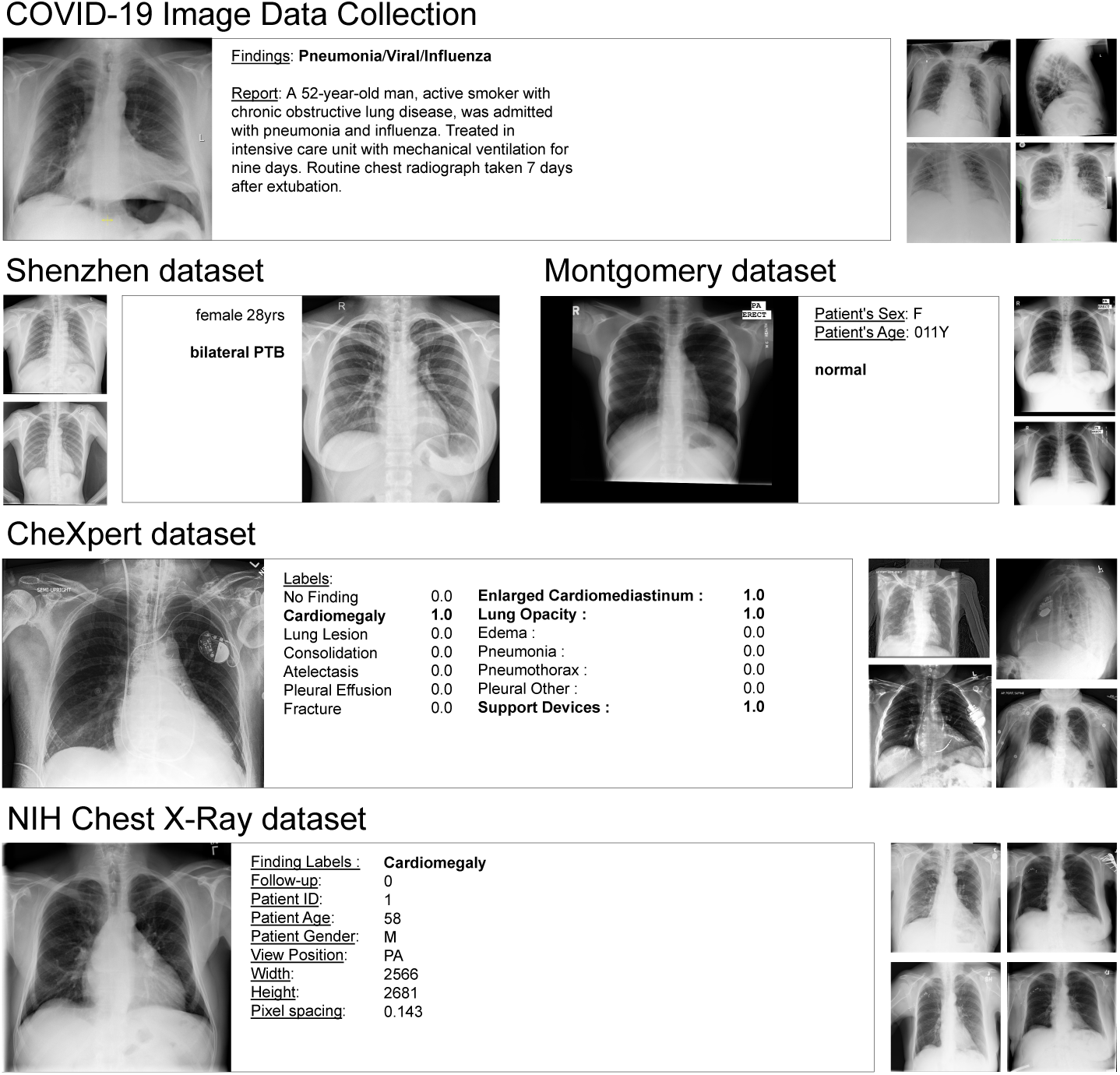
Samples and metadata from datasets used in this work. Data includes COVID-19 Image Data Collection [46], Montgomery and Shenzhen datasets [47], CheXpert dataset [32] and NIH Chest X-ray dataset [48].

Montgomery and Shenzhen datasets were obtained from medical centers in Montgomery County, MD, USA, and Shenzhen, China, and released by the US National Library of Medicine [47]. We combine them and obtain a dataset suitable for binary classification, with images labeled normal (healthy) or abnormal (tuberculosis). There are 406 cases with ”healthy” labels and 394 cases with ”tuberculosis detected” labels. Again, we randomly split the set to obtain the train/test set in proportions 80%/20%

The CheXpert dataset [32] is a large-scale chest X-ray dataset containing 224,316 images from 65,240 patients. We selected 11 pathologies with the largest number of positive cases available, resulting in 223,413 images in the training set. To every one of these cases, we assigned a positive label if the pathology was detected and a negative label otherwise. The corresponding test set is much smaller, with the exact numbers shown in Table 1.

**Table 1:**
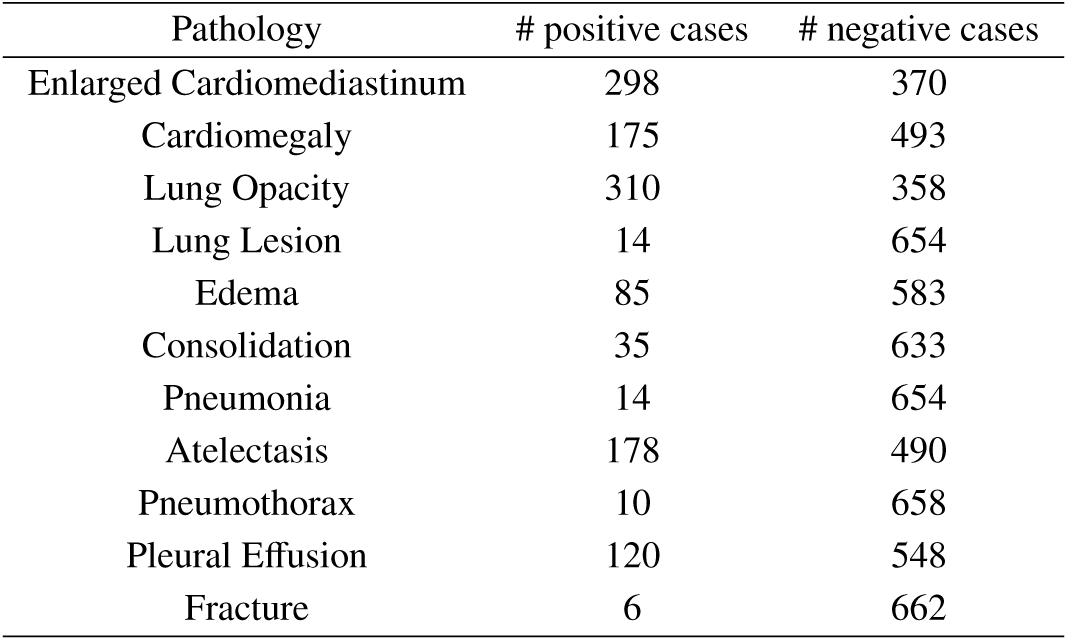
Number of examples available for selected pathologies in the test split of CheXpert dataset.

This NIH Chest X-ray dataset [48] contains 112,120 chest X-ray images from 30,805 patients. We sampled nine pathologies with the largest number of positive cases available, resulting in 86,524 training images. Every case with confirmed pathology was assigned a positive label, and a negative label was set for all negative or uncertain cases. We sampled the same pathologies from the test set, resulting in numbers in Table 2.

**Table 2:** Number of examples available for selected pathologies in the test split of NIH Chest X-ray dataset.

| Pathology | # positive cases | # negative cases |
| --- | --- | --- |
| Atelectasis | 3219 | 33788 |
| Cardiomegaly | 1069 | 35998 |
| Consolidation | 1825 | 35252 |
| Effusion | 4658 | 32409 |
| Infiltration | 6112 | 30955 |
| Mass | 1748 | 35319 |
| Nodule | 1623 | 35444 |
| Pleural Thickening | 1143 | 35924 |
| Pneumothorax | 2665 | 34402 |

### 2.6 Classification tasks for the generalized metric learning model

We chose two binary classification target tasks to evaluate our baseline selection of off-the-shelf feature encoders and adapted classifiers: COVID-19 and tuberculosis diagnosis. COVID-19 recognition task was composed from COVID-19 image data collection. Data for the tuberculosis recognition task was provided by Montgomery and Shenzhen datasets. In both cases the support set was randomly sampled from the train split, while the whole test set was used for evaluation.

For the evaluation of the model with domain-adapted feature extractors, we used a much larger set of target tasks to improve our ability to detect small effects of self-supervised fine-tuning. At first, we utilized binary classification tasks drawn from the CheXpert train dataset to see if the fine-tuning resulted in any few-shot classification improvement within the source dataset. We follow [29] and call this setting *in-distribution* (ID) evaluation. Next, we composed the *out-of-distribution* (OOD) set of binary classification tasks from NIH Chest X-ray dataset. This allowed us to assess the model generalization ability and transferability of knowledge learned on ID dataset.

In both cases (ID and OOD evaluations) we measured a relative mean change of AUROC achieved with feature extractors before and after domain adaptation. The statistical significance of the results was assessed using a paired two-tailed *t*-test with the null hypothesis that domain adaptation has no impact on performance. *p*-values less than 0.05 were considered statistically significant.

## 3 Results

The results of our experiments include the performance assessment of several variants of the model, in different few-shot settings. We present the comparison of model efficacy with the incorporation of general and domain-specific feature extractors, including our adapted versions of DINO-ViT-B/8, ViT-MAE-B and ResNet-50. We examine the performance of three described classifiers with the respect to the support data imbalance ratio. Lastly, we give a detailed analysis of the effectiveness of our attempt of self-supervised domain adaptation of DINO-ViT-B/8, ViT-MAE-B and ResNet-50 models.

### 3.1 General feature extractors

The results of the evaluation of general feature extractors for COVID-19 recognition are described in Table 3. ViTs outperformed CNNs in almost every scenario, with DINO-ViT-B/8 being the most effective. DINOv2 fell behind DINO-ViT and did not outperform ViT-MAE in most cases. Of the CNNs, ResNet-50 proved to be the most reliable, with ConvNeXt falling behind slightly and DenseNet-121 trailing them both significantly. Table 4 shows a similar comparison for the tuberculosis classification, but this time the results are very similar to the ones observed for COVID-19 task.

**Table 3:** Evaluation of general feature extractors for COVID-19 recognition task. Top 3 results measured by mean AUROC for each number of examples per class are marked in bold.

| Encoder | Classifier | Examples per class |  |  |  |
| --- | --- | --- | --- | --- | --- |
|  |  | 5 | 10 | 25 | 50 |
| ResNet-50 | k-NN | 0.578 | 0.615 | 0.654 | 0.681 |
|  | NC | 0.586 | 0.607 | 0.652 | 0.681 |
|  | NCA+k-NN | 0.565 | 0.606 | 0.652 | 0.686 |
| DenseNet-121 | k-NN | 0.519 | 0.528 | 0.545 | 0.561 |
|  | NC | 0.525 | 0.533 | 0.556 | 0.568 |
|  | NCA+k-NN | 0.511 | 0.527 | 0.540 | 0.555 |
| ConvNeXT-XL | k-NN | 0.566 | 0.587 | 0.612 | 0.627 |
|  | NC | 0.579 | 0.609 | 0.645 | 0.667 |
|  | NCA+k-NN | 0.565 | 0.588 | 0.631 | 0.656 |
| DINO-ViT-B/8 | k-NN | 0.591 | 0.617 | 0.667 | 0.706 |
|  | NC | **0.614** | **0.658** | **0.715** | **0.758** |
|  | NCA+k-NN | 0.592 | 0.618 | 0.669 | 0.706 |
| DINOv2-ViT-B/14 | k-NN | 0.581 | 0.610 | 0.665 | 0.693 |
|  | NC | **0.594** | 0.613 | **0.684** | 0.709 |
|  | NCA+k-NN | 0.585 | 0.602 | 0.663 | 0.695 |
| ViT-MAE-B | k-NN | 0.591 | **0.623** | 0.670 | **0.712** |
|  | NC | **0.599** | **0.656** | **0.700** | 0.737 |
|  | NCA+k-NN | 0.581 | 0.603 | 0.671 | 0.711 |

**Table 4:** Evaluation of general feature extractors for tuberculosis recognition task. Top 3 results measured by mean AUROC for each number of examples per class are marked in bold.

| Encoder | Classifier | Examples per class |  |  |  |
| --- | --- | --- | --- | --- | --- |
|  |  | 5 | 10 | 25 | 50 |
| ResNet-50 | k-NN | 0.630 | 0.672 | 0.719 | 0.752 |
|  | NC | 0.638 | 0.685 | 0.734 | 0.767 |
|  | NCA+k-NN | 0.626 | 0.685 | 0.748 | 0.786 |
| DenseNet-121 | k-NN | 0.575 | 0.600 | 0.639 | 0.657 |
|  | NC | 0.584 | 0.612 | 0.650 | 0.669 |
|  | NCA+k-NN | 0.544 | 0.579 | 0.623 | 0.659 |
| ConvNeXT-XL | k-NN | 0.639 | 0.674 | 0.709 | 0.730 |
|  | NC | 0.650 | 0.695 | 0.738 | 0.758 |
|  | NCA+k-NN | 0.629 | 0.684 | 0.734 | 0.766 |
| DINO-ViT-B/8 | k-NN | 0.739 | 0.776 | 0.807 | **0.834** |
|  | NC | **0.757** | **0.789** | **0.825** | **0.841** |
|  | NCA+k-NN | **0.739** | **0.782** | **0.817** | 0.831 |
| DINOv2-ViT-B/14 | k-NN | 0.729 | 0.778 | 0.808 | 0.819 |
|  | NC | **0.752** | **0.791** | **0.815** | 0.822 |
|  | NCA+k-NN | 0.733 | 0.771 | 0.805 | 0.823 |
| ViT-MAE-B | k-NN | 0.719 | 0.762 | 0.797 | 0.815 |
|  | NC | 0.718 | 0.768 | 0.812 | 0.827 |
|  | NCA+k-NN | 0.679 | 0.751 | 0.810 | **0.842** |

### 3.2 Domain-specific feature extractors

The Table 5 shows the comparison of performance achieved with domain-specific feature extractors. The REMEDIS model stands out in this task, outperforming our adapted DINO-ViT-Xray model by a large margin, with every combination of classifier and few-shot setting. ViT-MAE-Xray model performed similarly to DINO-ViT-Xray. Domain-specific CNNs performed relatively poorly compared to ViTs. Among these architectures, ResNet-50-RIN performed marginally better than others.

**Table 5:**
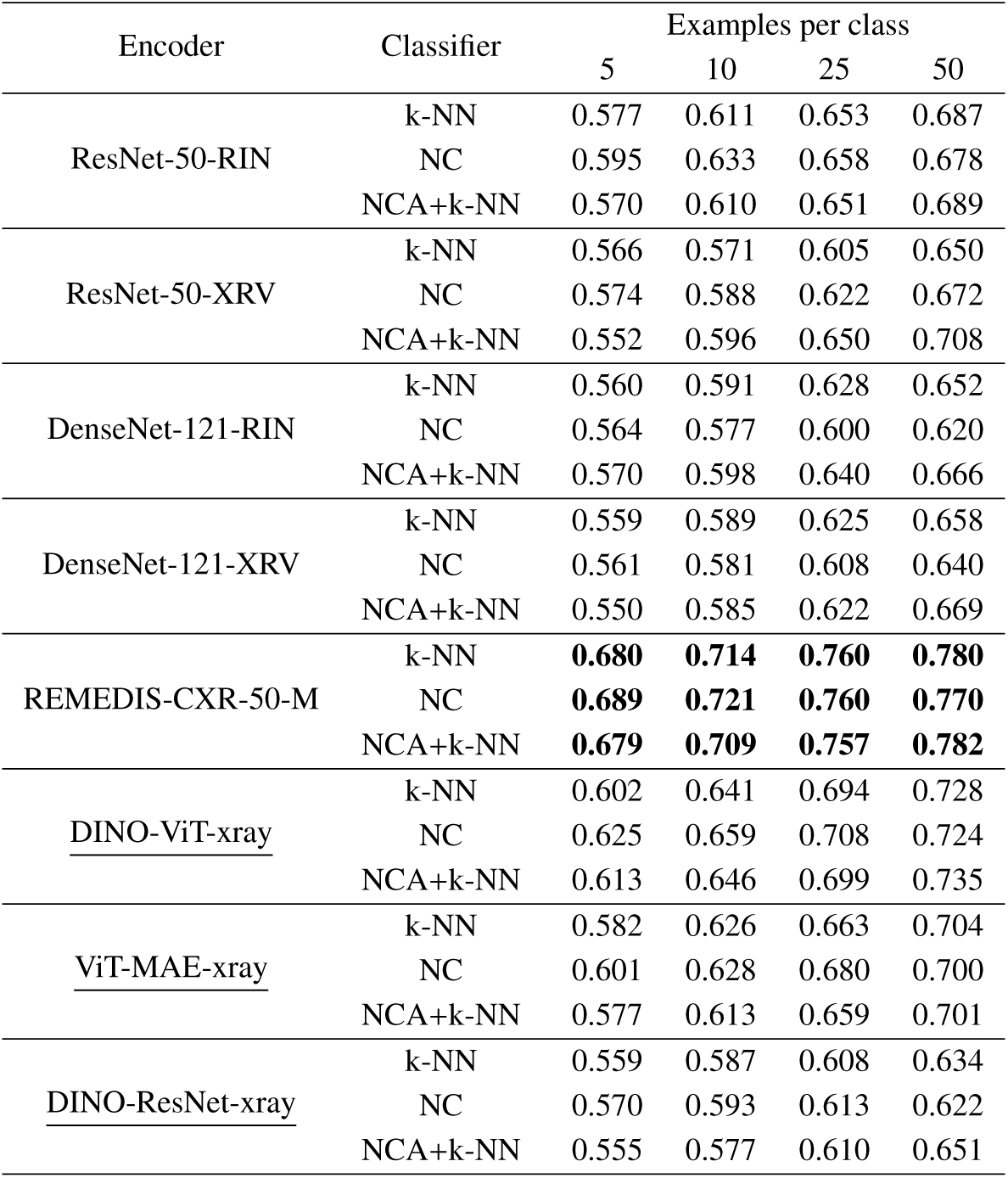
Evaluation of domain-specific feature extractors for COVID-19 recognition task. Models adapted by ourselves are underlined. Top 3 results measured by mean AUROC for each number of examples per class are marked in bold.

Different observations were made in the tuberculosis recognition task (Table 6), with supervised CNNs, ResNet-50-XRV and DenseNet-121-XRV, noting the highest AUROC across all domain-specific feature extractors. CNNs with RIN weights, however, performed worse with NC classifier, and the only improvement over their general versions is noted with k-NN and NCA+k-NN classifiaction heads. Our adapted DINO-ResNet-Xray performs very similar to the ResNet-50-RIN model, however the efficacy rises slightly with the NC classifier. The model also outperforms the adapted ViT-MAE-Xray. REMEDIS extractor performs much better than the RIN version of ResNet, however still stays far behind ResNet-50-XRV. DINO-ViT notes similar, yet slightly better efficacy.

**Table 6:**
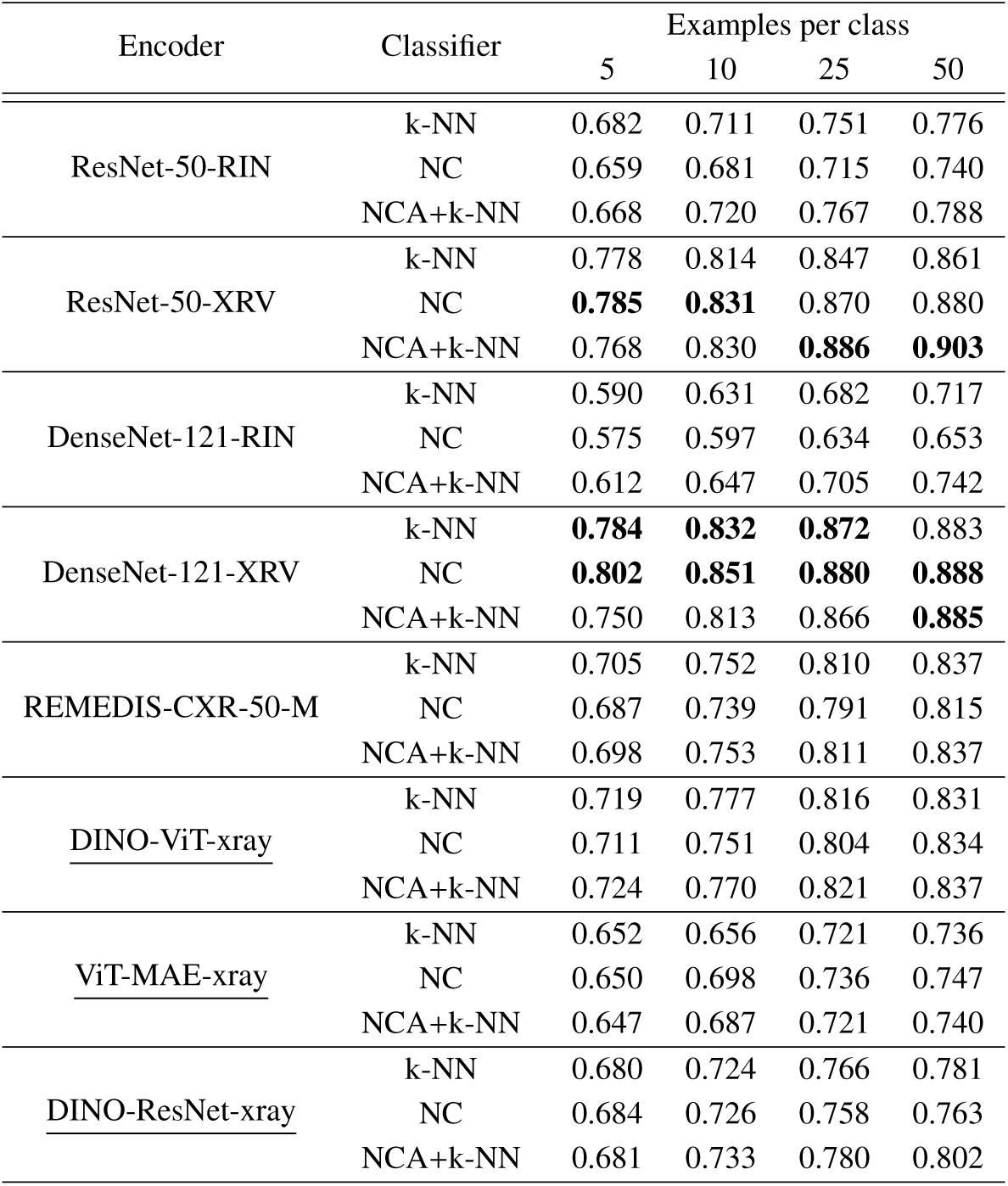
Evaluation of domain-specific feature extractors for tuberculosis recognition task. Models adapted by ourselves are underlined. Top 3 results measured by mean AUROC for each number of examples per class are marked in bold.

An additional comparison of general and domain-specific versions of DenseNet-121 and ResNet-50 is shown on Fig 3. The observations further emphasise relatively low performance of the base DenseNet-121, and superiority of REMEDIS in COVID-19 task. It is also clearly seen that XRV models are notably more effective than both RIN and REMEDIS models in tuberculosis classification task.

**Figure 3:**
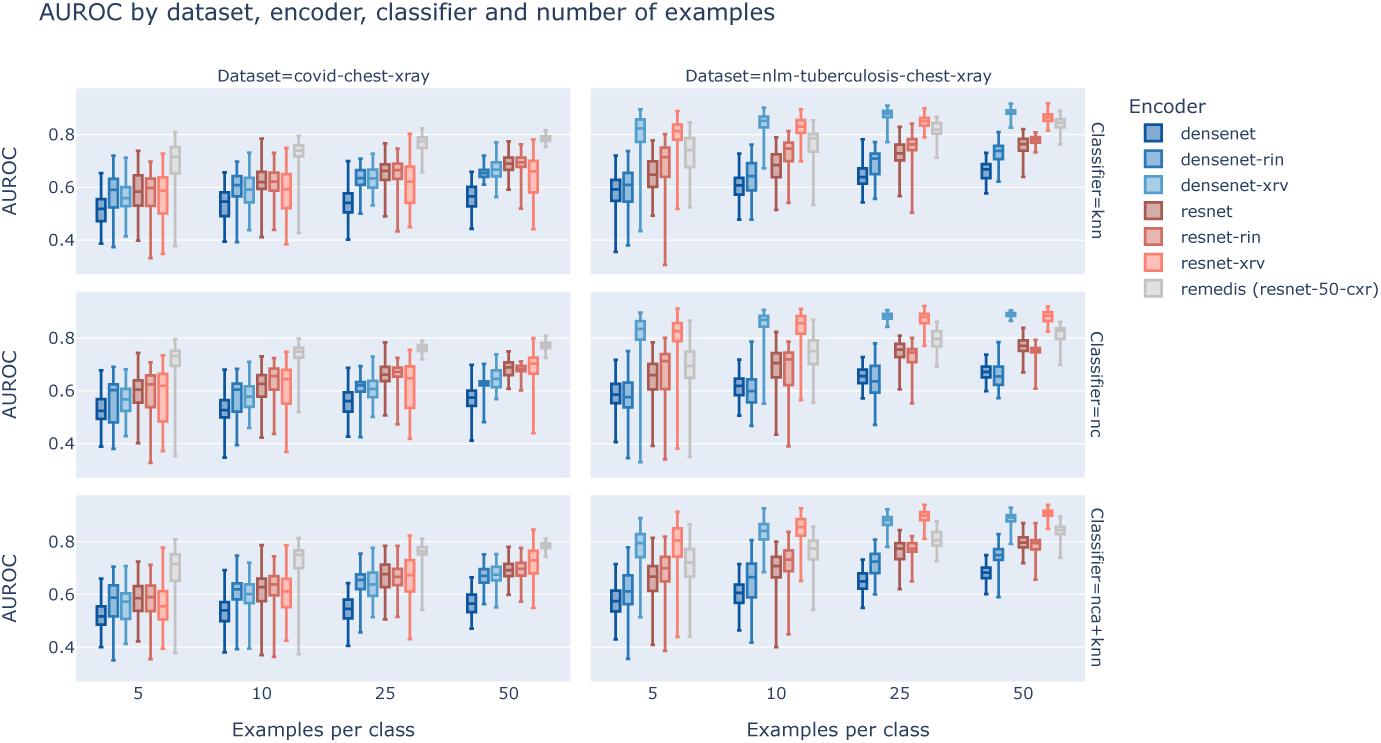
The comparison of general vs domain-specific feature extractors performance. The evaluation was done on COVID-19 and tuberculosis recognition tasks, with different combinations of classifier head and few-show setting.

We measured the relative change of mean AUROC after the domain adaptation of feature extractors. Results of ID evaluation are presented in Table 7. For ViT-MAE-B and DINO-ViT-B/8 we compared our self-supervised domain-adapted models with their base versions. DINO-ResNet-50 was juxtaposed with ResNet-50-XRV instead, to compare the efficacy of supervised and self-supervised domain-adaptation methods.

**Table 7:**
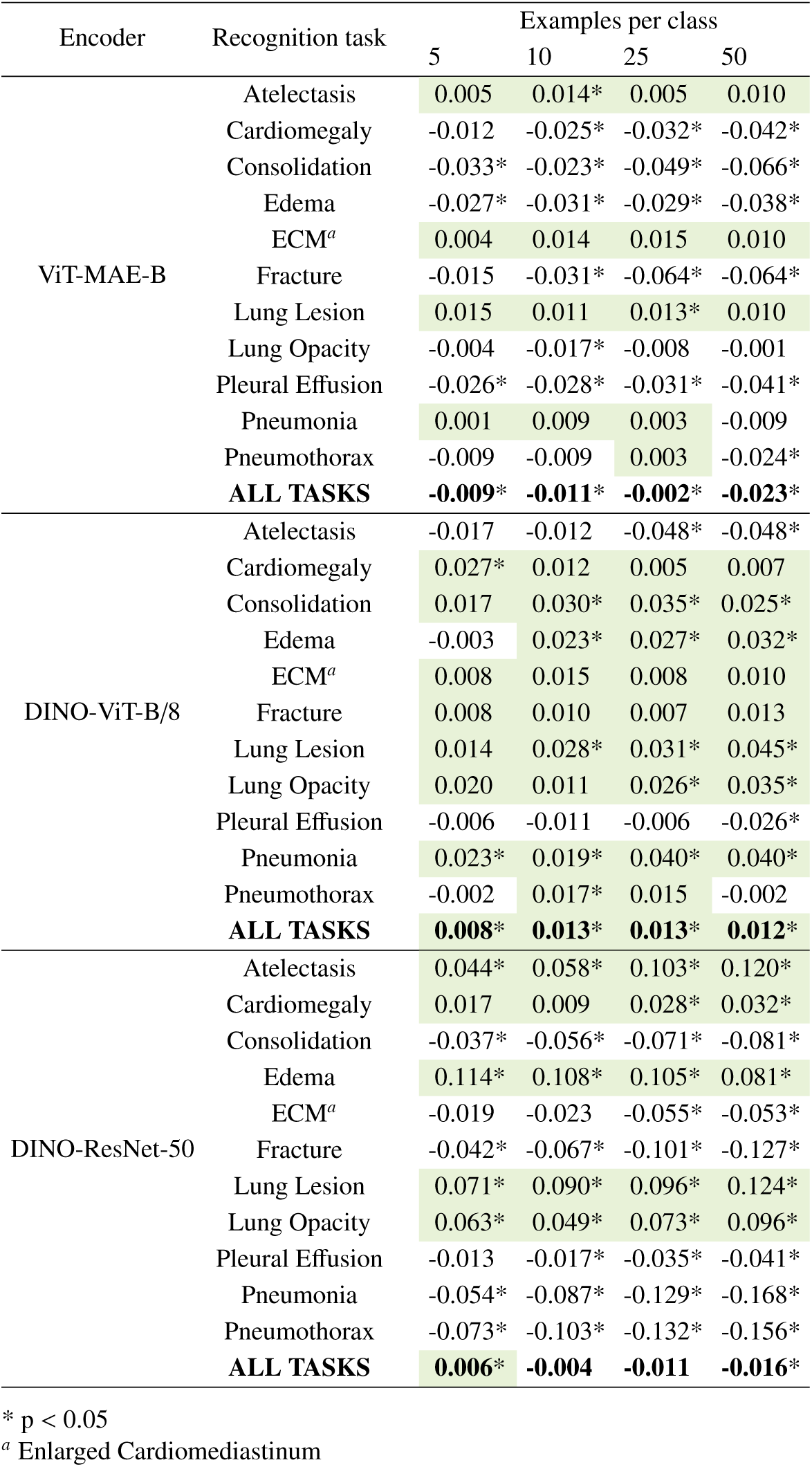
Relative mean AUROC change in ID tasks after domain adaptation of the encoder. In case of ResNet-50, the comparison was made with the relation to XRV model instead. Evaluation was performed on ID tasks sampled from CheXpert dataset. Positive changes are marked green. Significant differences were marked with stars.

Fine-tuning ViT-MAE-B improved the model performance (by mean AUROC) in 36% of tasks. Fine-tuning DINO-ViT-B/8 resulted in improvement in 75% of tasks, and fine-tuned DINO-ResNet-50 notes performance better than ResNet-50-XRV in 45% of cases. Averaging AUROC change across all tasks, DiNO-ViT performance increased by 0.0115, ViT-MAE performance decreased by 0.0146 and ResNet performance decreased by 0.0059.

Results of OOD evaluation are presented in Table 8. To sum it up, fine-tuning ViT-MAE-B improved the model performance in 22% of tasks, fine-tuning DINO-ViT-B/8 resulted in improvement in 6% of tasks, and fine-tuned DINO-ResNet-50 notes no performance improvement in any case. On average, DiNO-ViT performance decresed by 0.0195, ViT-MAE performance decreased by 0.0097 and ResNet performance decreased by 0.1195.

**Table 8:** Relative mean AUROC change in OOD tasks after domain adaptation of the encoder. In case of ResNet-50, the comparison was made with the relation to XRV model instead. Evaluation was performed on OOD tasks sampled from NIH Chext xray dataset. Positive changes are marked green. Significant differences were marked with stars.

| Encoder | Recognition task | Examples per class |  |  |  |
| --- | --- | --- | --- | --- | --- |
|  |  | 5 | 10 | 25 | 50 |
| ViT-MAE-B | Atelectasis | -0.014* | -0.018* | -0.017* | -0.017* |
|  | Cardiomegaly | 0.008 | 0.011* | 0.001 | 0.010* |
|  | Consolidation | -0.026* | -0.030* | -0.027* | -0.022* |
|  | Effusion | -0.011* | -0.006 | -0.012* | -0.014* |
|  | Infiltration | 0.008 | -0.001 | 0.005 | -0.004 |
|  | Mass | 0.004 | -0.010 | -0.009 | -0.013* |
|  | Nodule | -0.009 | 0.006 | -0.004 | -0.009 |
|  | Pleural Thickening | -0.003 | -0.003 | -0.005 | -0.005 |
|  | Pneumothorax | -0.013* | -0.026* | -0.031* | -0.046* |
|  | **ALL TASKS** | **-0.006\*** | **-0.086\*** | **-0.011\*** | **-0.013\*** |
| DINO-ViT-B/8 | Atelectasis | -0.018* | -0.031* | -0.027* | -0.029* |
|  | Cardiomegaly | -0.019* | -0.008 | -0.033* | -0.012* |
|  | Consolidation | -0.013 | -0.003 | -0.001 | -0.001 |
|  | Effusion | -0.045* | -0.048* | -0.051* | -0.046* |
|  | Infiltration | -0.007 | -0.004 | -0.007 | -0.025* |
|  | Mass | 0.001 | -0.004 | -0.014* | -0.013* |
|  | Nodule | -0.005 | -0.033* | -0.030* | -0.039* |
|  | Pleural Thickening | -0.006 | 0.003 | -0.001 | -0.003 |
|  | Pneumothorax | -0.021* | -0.030* | -0.048* | -0.033* |
|  | **ALL TASKS** | **-0.015\*** | **-0.018\*** | **-0.024\*** | **-0.022\*** |
| DINO-ResNet-50 | Atelectasis | -0.047* | -0.062* | -0.075* | -0.080* |
|  | Cardiomegaly | -0.136* | -0.162* | -0.191* | -0.206* |
|  | Consolidation | -0.055* | -0.071* | -0.068* | -0.063* |
|  | Effusion | -0.133* | -0.134* | -0.137* | -0.135* |
|  | Infiltration | -0.045* | -0.047* | -0.081* | -0.092* |
|  | Mass | -0.144* | -0.172* | -0.200* | -0.221* |
|  | Nodule | -0.059* | -0.083* | -0.114* | -0.123* |
|  | Pleural Thickening | -0.076* | -0.085* | -0.105* | -0.098* |
|  | Pneumothorax | -0.162* | -0.206* | -0.216* | -0.217* |
|  | **ALL TASKS** | **-0.095\*** | **-0.114\*** | **-0.132\*** | **-0.137\*** |

\* $p < 0.05$

### 3.3 Classifiers overview

The examination of model performance with respect to different support set imbalance ratios in COVID-19 and tuberculosis recognition tasks are shown on Figs 4 and 5. The results indicate that NC classifier is not only highly robust to the imbalance ratio, but in many cases achieves the best overall performance even when data is balanced, in every *k*-shot setting.

**Figure 4:**
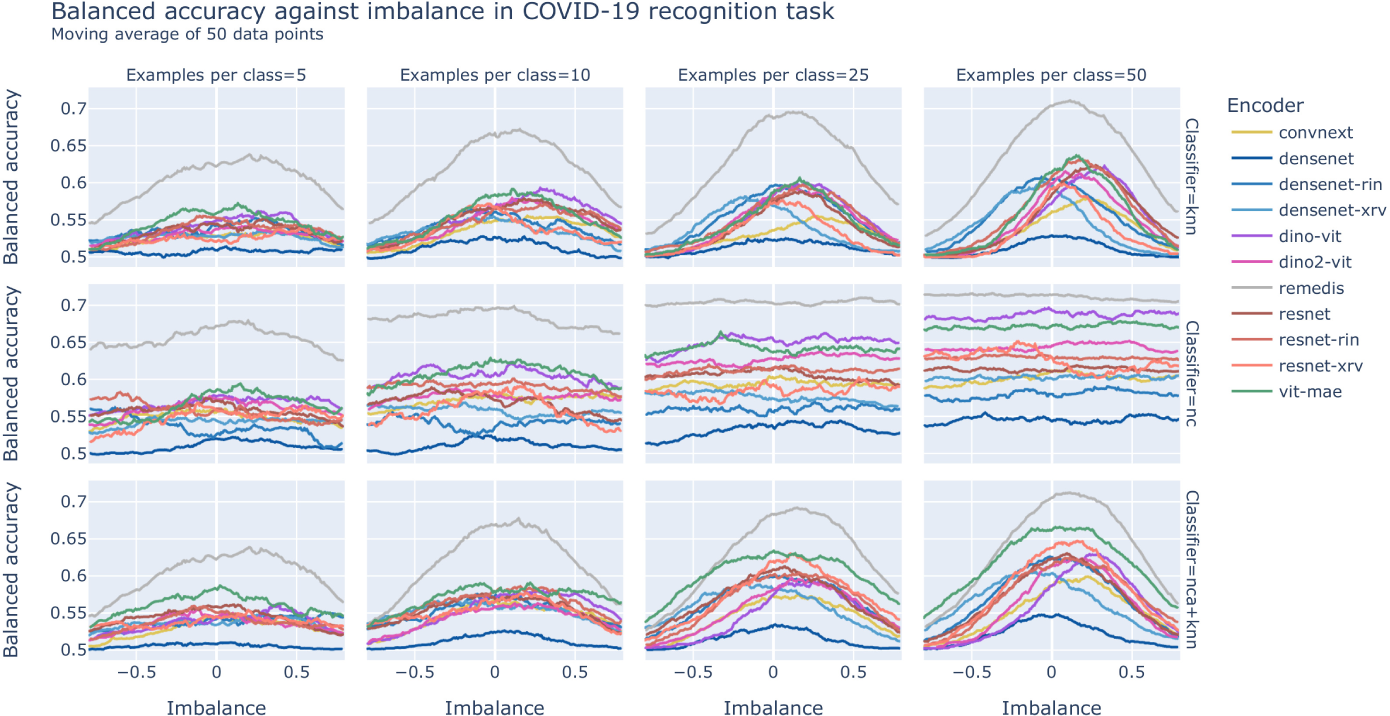
Balanced accuracy score in COVID-19 recognition task in different data imbalance conditions. Performance with respect to encoder, classifier, support data imbalance and number of examples per class.

**Figure 5:**
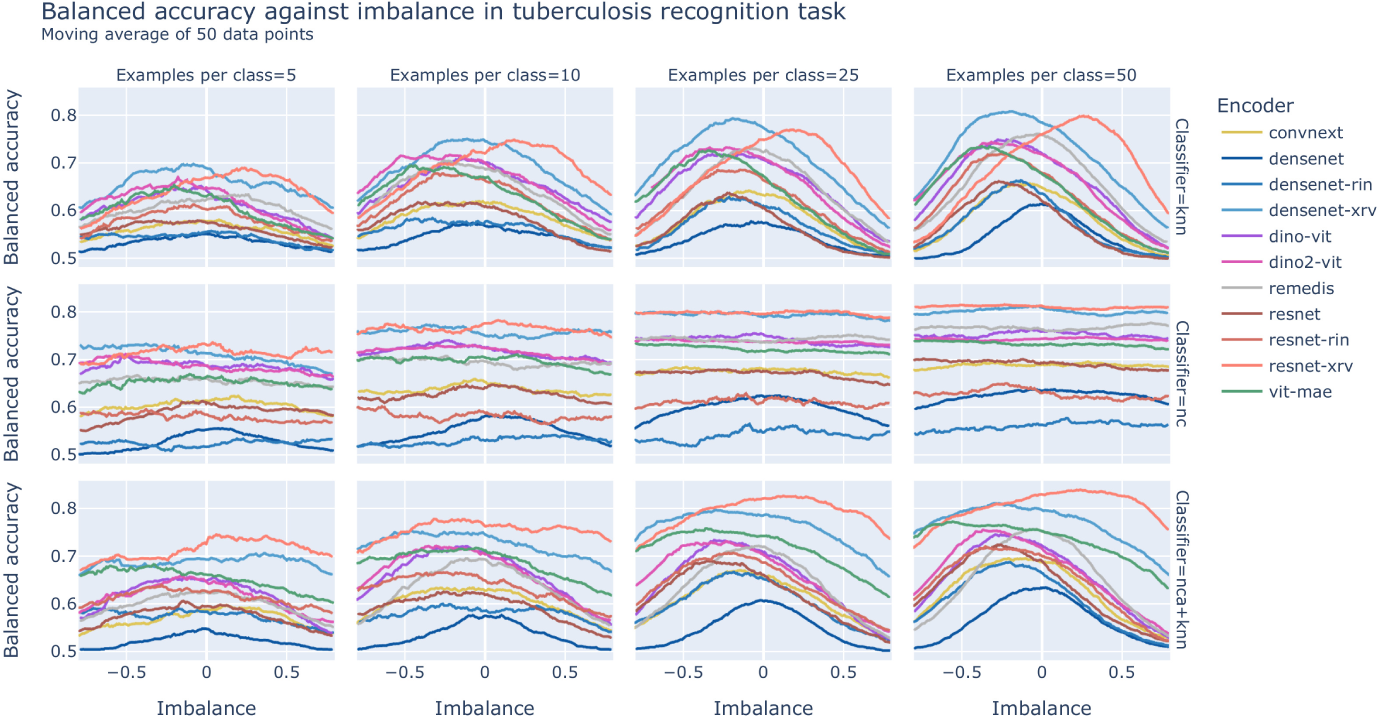
Balanced accuracy score in tuberculosis recognition task in different data imbalance conditions. Performance with respect to encoder, classifier, support data imbalance and number of examples per class.

## 4 Conclusion and discussions

Our work provides a systematic evaluation of the metric learning approach in several few-shot medical image classification tasks. Our experiments have shown that the nearest centroid algorithm is a much more reliable choice as a classification head than k-NN, outperforming the latter in almost every few-shot scenario. We further conclude that both DINO and MAE vision transformers may be a good selection as feature extractors in metric learning models, outperforming general-purpose CNNs by a large margin. The recently updated second version of DINO did not perform better than its baseline in our setting.

The performance of medical-trained CNNs varies, although the best results are seen when the model training domain is closer to the target tasks (XRV models generally outperform RIN). Domain-specific REMEDIS model achieves remarkable results in COVID-19 detection task and is comparable to the XRV and RIN models in tuberculosis recognition. Other self-supervised models, however, noted varying changes in performance after fine-tuning them on the source dataset. This relative performance became straightly negative when the model was applied to out-of-distribution tasks. This leads us to a counter-intuitive conclusion that some self-supervised models may have a greater ability to extract important features from medical data if trained in a natural domain, as we observed with DINO and MAE.

While in our experiments we did not come close to the state-of-the-art classification performance noted by non-few-shot learning models, the simplicity of the presented approach easily allows for further improvement of the pipeline through mixing in other few-shot techniques, such as meta-learning or ensemble methods.

### 4.1 Di**ff**erent model variants comparison

The superiority of REMEDIS model in COVID-19 classification task (Table 5) is not surprising, as this framework is a recent state-of-the-art solution carved purposely to solve medical classification problems, and the CXR-50 version of this model was trained on chest X-rays. Its high performance suggests that relatively simple and long-established CNN architectures can achieve outstanding performance in medical imaging and the training strategy is the key factor in that matter.

Aside REMEDIS, ViT models proved to be the best choice for feature extractors in this task, outperforming even domain-specific RIN and XRV models in every case. Surprisingly, the DINOv2-ViT model did not outperform the base DINO-ViT as might have been expected [36]. ViT-MAE model noted slightly lower performance than DINO, which goes on par with conclusions from [29, 24] stating that contrastive self-supervised approaches work better than image-reconstruction-based ones. The CNNs’ performance was observed to be comparable, except the base DenseNet-121 which fell behind noticeably (this is also seen clearly on Figs **??** and **??**).

In the case of tuberculosis classification, the best performance was obtained through the incorporation of XRV models, with ViTs being close behind and outperforming RIN models. Again, DINOv2 performed slightly worse than the base DINO. This time REMEDIS model achieved performance similar to ViT models. What is also interesting is that in the tuberculosis task the application of DenseNet-121-RIN results in worse performance than the base DenseNet-121. This suggests that in some cases models trained on natural images may have a greater ability to extract important features from X-ray data than the ones trained on medical, but not X-ray images.

Figure ??, shows that in a difficult task, such as COVID-19 recognition, the performance of CNN model can be improved by training it on medical data in a supervised way, no matter if it is in-domain (XRV models) or out-of-domain (RIN models). For the easier task, however, the advantages of in-domain training start to become clear, with XRV models greatly outperforming RIN ones.

To compare our results to the state-of-the-art, we note that Zhang et al. [49] reports the performance of the proposed COVID-19 screening method as 0.952 AUROC. Next, Tartaglione et al. [50] report the COVID-19 recognition performance as high as 1.0 AUROC, however, the specificity of the proposed solution is only 0.20. Interestingly, in a work of Shorfuzzaman and Hossain [51] the authors report 0.975 AUROC for 3-way 10-shot classification of COVID-19 and pneumonia with the use of contrastive learning and Siamese network. This highlights the potential of self-supervised learning in medical few-shot imaging. It also indicates that our approach still needs refinement to compete with the most effective methods established for this task. Our model with REMEDIS-CXR-50-M feature extractor achieved 0.721 AUROC in the 2-way 10-shot COVID-19 classification scenario and 0.782 AUROC in 2-way 50-shot setting.

In the work of Saif et al. [52] the AUROC in tuberculosis classification is set at 0.997 for the Montgomery dataset and 0.981 for the Shenzhen dataset. This was achieved through ensemble voting of different handcrafted and deep-learned features with data augmentation. Cahndra et al.[53] achieved 0.95 AUROC on Montgomery and 0.99 AUROC for Shenzhen set with the application of hierarchical feature extraction. Lastly, Rajamaran et al. [54] reports 0.954 AUROC on Shenzhen and 0.964 AUROC on Montgomery datasets. These results were achieved through the bone suppression technique, and the baseline performance without it is stated to be 0.899 and 0.857 AUROC respectively. This goes on par with our results, as we achieved 0.903 AUROC on these datasets combined in the more difficult 50-shot setting. Some of the works mentioned above mark the possible directions to improve our work in the future, either through introducing ensemble voting or incorporating bone suppression. As our approach is simple in conception, both of these methods would be relatively straightforward to implement within our framework.

### 4.2 Self-supervised domain adaptation

The results of self-supervised domain adaptation experiments suggest that domain-specific feature extractors are sometimes less effective than general-purpose ones. While the REMEDIS model shows great performance in COVID-19 and tuberculosis recognition tasks, our own attempts to adapt DINO and MAE frameworks similarly were not that successful. It can be said that while their average performance on ID tasks improved (Table 7), this is not the whole picture. In cases such as pneumothorax or edema recognition, there were many scenarios where the performance did not change significantly or even straightly decreased. In the worst case, in the pleural effusion detection task, the measured AUROC dropped in every scenario across all adapted feature extractors. On the other hand, there is an example of a lung lesion recognition task where the performance improved across all scenarios. This inconsistency of results is most notably seen after ResNet fine-tuning when the difference in the performance of the adapted model varied from almost −0.17 to 0.12 AUROC. It shows that it is difficult to indicate which feature extractor is the best for domain adaptation based only on mean performance change, and that the stability of results across many tasks must also be included in the analysis.

The results of the out-of-distribution evaluation (Table 8) are even more discouraging, as there is only one case (ViT-MAE cardiomegaly recognition) where the performance consistently improved. It suggests that the knowledge learned during the self-supervised training sometimes helps with classification on the source dataset, but is not always transferable to OOD tasks. While the reports from [40], [41] and [29] confirm the effectiveness of self-supervised domain adaptation, which we confirm for REMEDIS model [29], our own experiments indicate that this rule should not be uniformly applied to every self-supervised framework. Next, [41] reports minor improvement in the performance after the adaptation of DINO ResNet-50 and DINO DeiT-S transformer [55], yet the gains were mostly within the range of measured standard deviation. While this still marks the domain adaptation as a valid direction for improving a model’s efficacy, our experiments show that the gains from self-supervised pre-training should not be taken for granted.

## Data Availability

COVID-19 Image Data Collection files are available at https://github.com/ieee8023/covid-chestxray-dataset Images from Montgomery and Shenzhen datasets are available at https://lhncbc.nlm.nih.gov/LHC-downloads/downloads.html#tuberculosis-image-data-sets Images from CheXpert dataset are available via the links at the datasets homepage: https://stanfordmlgroup.github.io/competitions/chexpert/ Images from NIH Chest X-Ray dataset are available at https://www.kaggle.com/datasets/nih-chest-xrays/data

https://stanfordmlgroup.github.io/competitions/chexpert/

https://lhncbc.nlm.nih.gov/LHC-downloads/downloads.html#tuberculosis-image-data-sets

https://github.com/ieee8023/covid-chestxray-dataset

## Footnotes

1 https://github.com/facebookresearch/dino

2 https://github.com/facebookresearch/mae

